# Urinary collagen-derived peptides as sensitive markers for bone resorption and bisphosphonate treatment in kidney transplant patients

**DOI:** 10.1101/2022.02.15.22270979

**Authors:** David Marx, Dany Anglicheau, Sophie Caillard, Bruno Moulin, Audrey Kochman, Harald Mischak, Martin Pejchinowski, Agnieszka Latosinska, Frank Bienaimé, Dominique Prié, Pierre Marquet, Peggy Perrin, Wilfried Gwinner, Jochen Metzger

## Abstract

Kidney transplant recipients (KTR) are at increased risk of fractures. Total urinary hydroxyproline excretion used to be a marker for bone resorption (BR) but faded into the background when more specific markers like Beta-CrossLaps (CTX) became available. Proteomic studies identified numerous hydroxyproline-containing urinary collagen peptides but their origin remains unknown. We followed the hypothesis that some of the urinary collagen peptides are associated with BR and are markers for pathophysiological changes in bone metabolism of KTR. Clinical and laboratory data including serum levels of CTX in 96 KTR from two French centers (Strasbourg, n=38; Paris-Necker, n=58) were correlated with the signal intensity of urinary peptides identified by capillary electrophoresis (CE) coupled to mass spectrometry (MS) and tandem-MS. The effect of oral bisphosphonates on urinary peptides was studied in an independent group of 11 KTR. Eighty-two urinary peptides were identified to be significantly correlated with serum CTX levels in both cohorts. Statistical association with parameters other than BR markers were not significant. Collagen α-1(I) chain (COL1A1) was the most frequently identified peptide source. COL1A1 peptides associated with BR were significantly more hydroxylated than those showing no association (55.9% versus 45,2%, p<0.0003 by a χ^2^-test). From the 82 urinary peptides correlated to CTX, 17 were significantly associated with bisphosphonate treatment. All of these 17 peptides showed a marked reduction in their excretion levels after 410 ± 344 days of bisphosphonate treatment compared to baseline levels. We studied the cleavage sites of these COL1A1 peptides and observed a signature of Cathepsin K and Matrix Metallopeptidase 9. This study provides strong evidence for the occurrence of collagen peptides in the urine of KTR that are associated with BR and that are sensitive to bisphosphonate treatment. Their assessment might become a valuable tool to monitor bone status in KTR.

## Introduction

Kidney transplant recipients (KTR) are at an increased risk of bone fractures (1–4). In the first year after renal transplantation, a drop in bone density of 7-9% on average occurs, which is in part attributed to a shift towards greater rates in bone resorption relative to bone formation (5). This excessive resorption befalls patients who already have a history of severe chronic kidney disease (CKD) and anomalies in bone metabolism summarized under the common term of CKD - mineral bone disease (CKD-MBD). KTR therefore keep a high incidence of fractures in the 5 years following transplantation (6). Although many factors that favor the persistence of increased bone resorption, such as treatment with corticosteroids, hyperparathyroidism, hyper/hypophosphatemia, hypomagnesemia, and vitamin D deficiency in the post-transplant context were identified (7), the pathophysiological and molecular mechanisms are not well understood.

Dual-energy X-ray absorptiometry commonly used for the study of bone density cannot reliably predict individual fracture risk in KTR(8). Bone biopsy is rarely performed given its invasive nature. Due to these reasons noninvasive monitoring with serological markers of formation and resorption is commonly used in KTR and includes osteocalcin, bone alkaline phosphatase, and amino-terminal and carboxy-terminal collagen-I crosslinks (NTX and CTX respectively). Most of these markers, however, also have a drawback: they are excreted by the kidneys and levels vary depending on renal function, the latter being quite variable in KTR(9).

Depending on the context and on the estimated risk of bone loss and fracture, KTR are often treated with a combination of one or more drugs including vitamin D, calcium supplements, bisphosphonates, denosumab or hormone replacement therapy. Still, many patients with rapid decline of bone mass are missed. Hence, there is a clear need for diagnostic markers to better identify KTR with rapid bone loss being at increased risk of bone fracture and who could benefit from these treatments.

In the 1960’s-1980’s, a vast number of medical references established an association between urinary collagen peptides and bone resorption (BR) both in humans and in animal models (4– 12). Essentially, these studies showed that there are peptides containing hydroxyproline (HyP) in the urine, which are in part derived from bone and are associated with BR. At the time of the discovery of these peptides, however, their amino acid sequences could not be determined. In line with this, total urinary HyP (including free and peptide-bound HyP) has been used as a marker of BR during several decades but urinary HyP peptides from other sources including food intake and skin reduced its diagnostic performances (10). Advances in tandem mass spectrometry (MS) have recently made it possible to better characterize these peptides including information on their AA sequence (11). More recently MS has tremendously improved analysis of urinary proteins and peptides. Studies have shown the wide variety of low molecular weight peptides in the urinary environment, even in healthy subjects(11–15). In particular, urine displays a large variety of collagen fragments, especially COL1A1. The detailed analysis of the AA sequences of these fragments reveals the richness in glycyl-prolyl-HyP (GPp) sequences(16). Hundreds of different fragments of this type have been identified, using both capillary electrophoresis (CE) and liquid chromatography as a separation technique (see for instance the supplemental files of reference(14)). One study compared low molecular weight peptides from blood and urine using CE coupled to MS (CE-MS) (17) and showed that the overlap was to a large extent made up of collagen peptides. Urine and blood levels of these peptides were found to be correlated to each other. However, the origin and biological significance of the vast majority of urinary collagen peptides is unknown.

In the current study, we hypothesized that urinary, bone-derived collagen peptides evidenced in the past for which the precise nature remained unknown, could now be precisely identified using MS and serve as source for new diagnostic markers for bone remodeling and response to therapy. As a first step into this direction, we study the low molecular weight urinary proteome of individuals from two cohorts of renal transplant patients and search for peptides whose quantification is correlated with the concentration of established bone remodeling markers. Peptides fitting this description are then characterized in terms of AA sequence, type of association with bone resorption, HyP content, relation to bone proteolytic activity and finally sensitivity of their urine levels to bisphosphonate therapy.

## Methods

### Patients and samples

The 96 KTR patients used in this study originate from two clinical centers in France and are a representative cohort of graft recipients in the first-year post-transplantation. The patient cohort from the University Hospital of Strasbourg consists of two subgroups. One group consists of 27 patients with various degrees of BK virus-related post-kidney transplantation complications from whom samples were taken at one single time point after kidney transplantation. The second group from this center are 11 KTR patients affected by bone loss due to pre-transplant osteopenia or osteoporosis with samples taken at two different time points namely before the initiation and after several months of treatment with bisphosphonates. A third KTR study cohort consisting of 58 patients monitored for serum BRM is derived from the Necker Hospital in Paris representing a cross-sectional group of patients with (n=29, 8 ABMR, 5 TCMR, 16 IFTA) and without (n=29) transplantation-related complications.

Bone metabolism is routinely monitored by serum CTX and BAP at the Univ. of Strasbourg and by serum CTX and osteocalcin at the Necker Hospital in Paris. Urine samples collected within a time range of less than 15 days from the time of BRM assessment were selected from the biobanks of the two hospital centers to perform proteomic analysis by CE-MS. Clinical and demographic characteristics for the KTR patients from the University Hospital of Strasbourg and the Necker Hospital in Paris are given in Table 1.

**Table 1.**
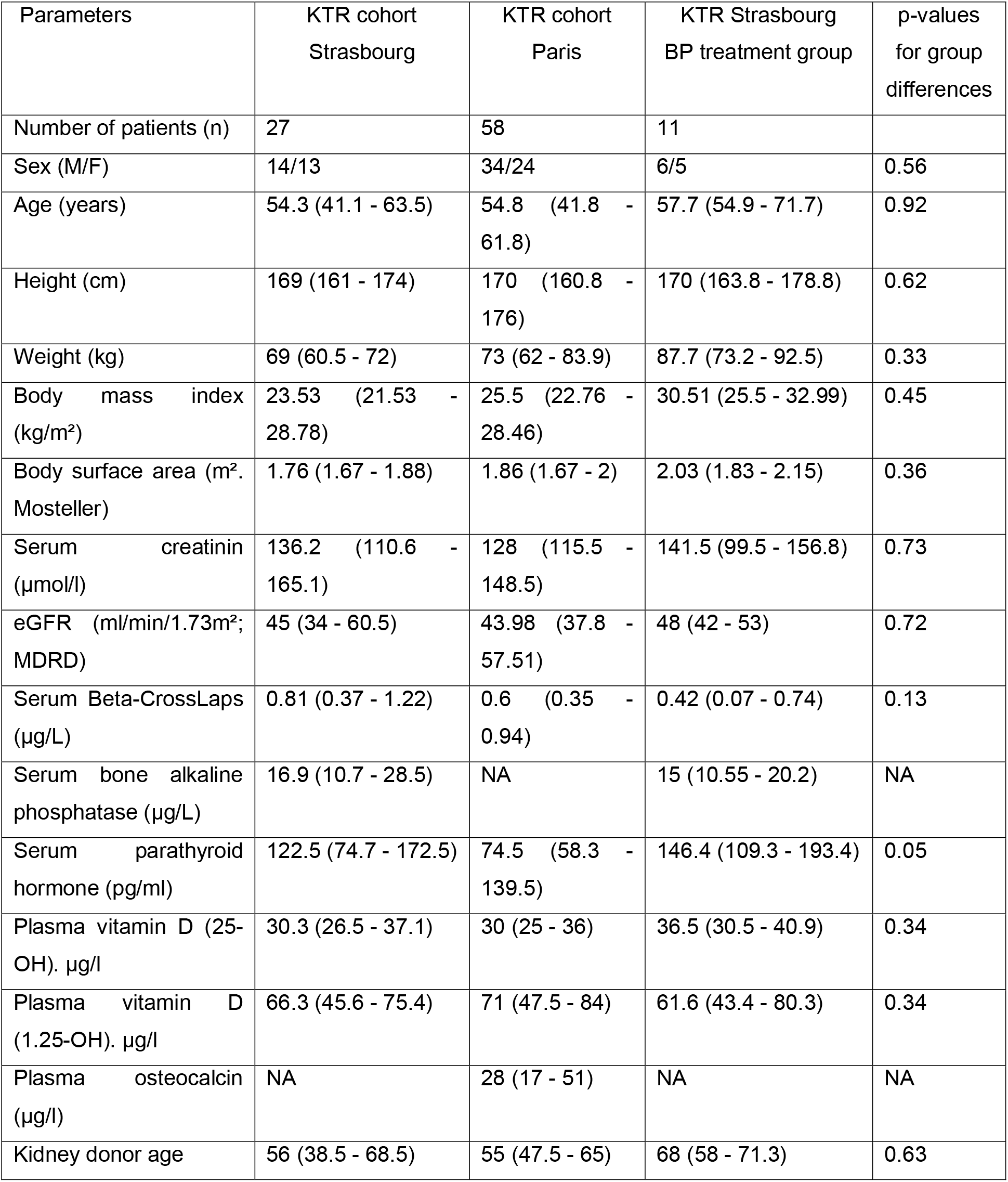
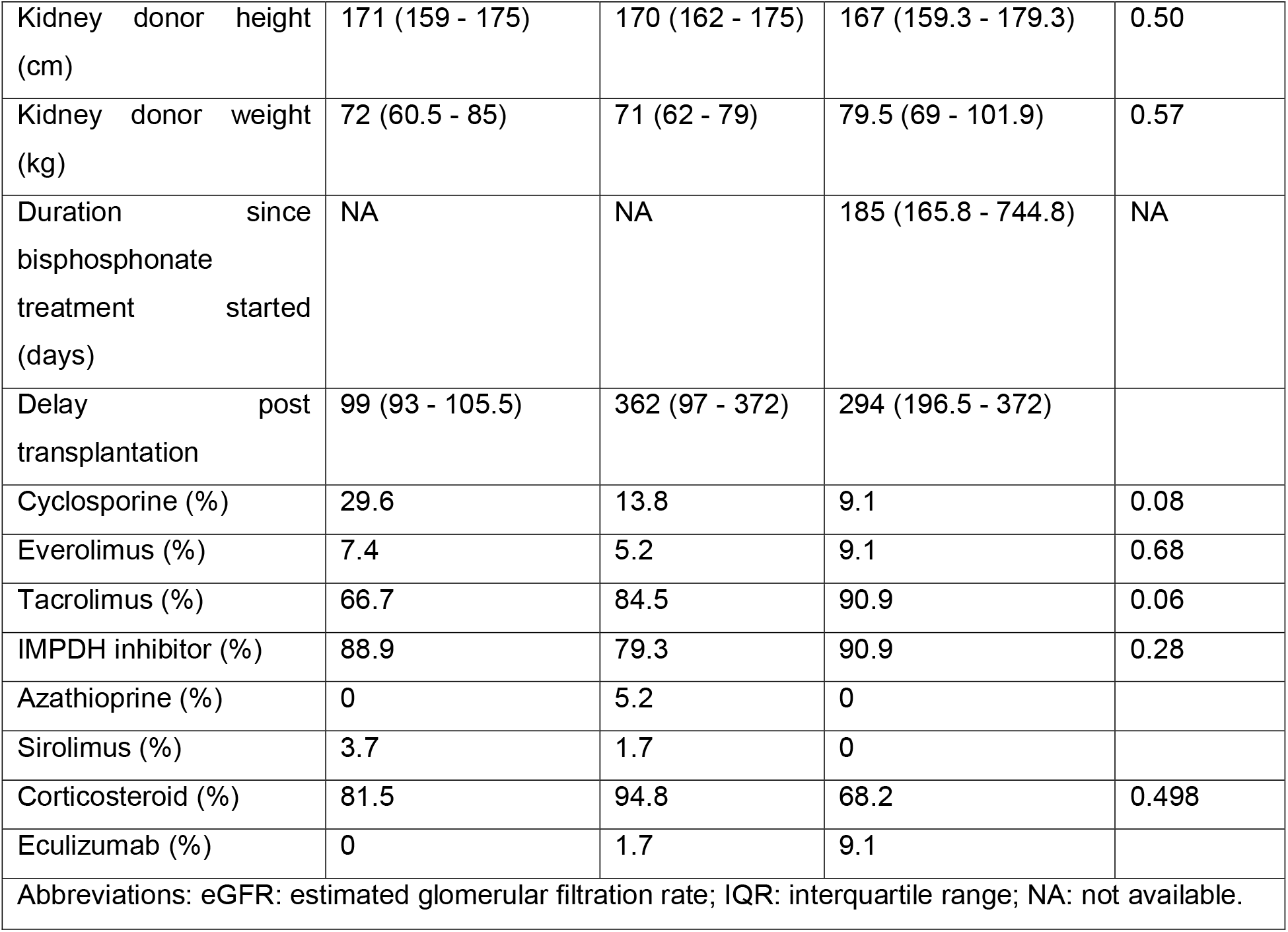
Clinical and demographic characteristics of the patients from the three study groups. Values are given as median and interquartile range in square brackets unless stated otherwise. In the bisphosphonate (BP) study group, treatments were assessed for each of the 22 time points of sample procurement.

All patients provided informed consent. The study was approved by the institutional review board of Strasbourg University Hospitals under the reference CPP-EST DC-2013-1990 for patients from Strasbourg. For patients from the Necker Hospital the study was approved by the Ethics Committee of Ile-de-France XI (#13016).

Spot urine samples were transferred to urine monovettes and immediately frozen without additives at -80 °C. Shipment to the central laboratory of the CE-MS conductor Mosaiques Diagnostics was carried out on dry ice in order to thaw them only once before CE-MS analysis.

### Immunoassays and laboratory tests

Demographic and clinical data was collected from each KTR and respective kidney allograft donor and included age, sex, height, weight, body surface area (using the Mosteller formula (18)), body mass index (kg/m²). Laboratory markers from the KTR included serum creatinine, estimated glomerular filtration rate (eGFR) by the MDRD formula (19), serum PTH, serum 25(OH) vitamin D and 1,25(OH)2 vitamin D as determined by the use of standard clinical laboratory methods. For patients from Strasbourg serum CTX-I and BAP were analyzed using the Automated Chemiluminescence Immunoassay (CLIA) (Crosslap®, Ostase®) on an iSYS automate (IDS-iSYS Multidiscipline Automated System). In patients from Paris CTX and osteocalcin were analyzed by the Elecsys® b-CrossLaps and the N-MID Osteocalcin serum assays respectively (Roche Diagnostics) on the Cobas® e411 analyzer.

### Urinary peptide analysis

Proteomic analysis of urine samples by CE-MS was performed as described elsewhere (14). Briefly, urine was prepared by 1:2-dilution in 2 M urea, 10 mM NH4OH and 0.02% SDS (pH 10.5), followed by ultracentrifugation through Centrisart devices (Sartorius, Germany) and desalting using PD10 columns (GE Healthcare, Sweden) to remove proteins above 20 kDa, as well as salts and lipids. Samples were then lyophilized, resuspended in HPLC-grade water to a protein concentration of 0.8 µg/µL and a 100 nL-aliquot injected into the CE-MS under constant flow and pressure conditions at a pH of 2.2 to ensure that all peptides are positively charged. CE-MS sample acquisition was performed using a P/ACE MDQ capillary electrophoresis system (Beckman Coulter, Fullerton, USA) on-line coupled to a Micro-TOF MS (Bruker Daltonic, Bremen, Germany). Prerequisite for coupling of the CE with the MS was that the ESI sprayer (Agilent Technologies, Palo Alto, USA) was grounded, the ion spray interface potential set between -4.0 and -4.5 kV and contact-close-relays were used to control MS data acquisition automatically by the CE. Spectra were accumulated every 3 s over a m/z range of 350 to 3000. For raw proteomic data processing, the in-house software MosaiquesVisu (20) was used to deconvolute mass spectral ion peaks representing identical molecules at different charge into single masses. MosaiquesVisu uses both isotopic distribution as well as conjugated masses for charge-state determination of peptides with charge states >1 and signal-to-noise ratios >4 observed in a minimum of 3 consecutive spectra. The software generates a list of peptides in the sample characterized by molecular mass, CE-migration time, and ion signal intensity, which were subsequently normalized by global and local linear regression using internal standard peptides of high abundance, low ion signal variability and FT-ICR MS-determined exact molecular mass ensuring inter-comparability across measurements (21). The normalized peptide profiles were deposited, matched, and annotated in a Microsoft SQL database, allowing further analysis and comparison of multiple samples. Peptides were considered identical within different samples, when mass deviation was lower than 50 ppm for peptides less than 2,000 Da or 75 ppm for larger peptides. After calibration, deviation of migration time was controlled to be < 0.45 min.

### Peptide sequence analysis

For amino acid sequence identification, CTX-associated urinary peptides derived from the CE-MS spectra based on their MS-detected molecular mass and characteristic migration time in the CE were searched against Mosaiques’ in-house human urinary peptide database containing sequences from both LC-and CE-MS/MS using a Dionex Ultimate 3000 RSLS nano flow system (Dionex, Camberly, UK) for LC- and a Beckman CE/Orbitrap Q Exactive plus combination for CE-MS/MS (Thermo Scientific, Waltham, MA) (22).

### In silico protease mapping

In silico protease maps were generated using Proteasix (available online at http://proteasix.org/), which allows retrieving protease/cleavage sites (CS) associations based on the MEROPS (https://www.ebi.ac.uk/merops/) and the SWISS-PROT database (http://www.uniprot.org/)(23). N- and C-terminal cleavage sites were retrieved for the sequenced BR-associated peptides, based on an alignment to the respective substrate sequence information. The association of proteases with their cleavage sites is based on octapeptide consensus sequences. A list of six thousand random CS sequences was used as reference to determine the specificity of prediction against false-positive associations. Only proteases/CS associations which were previously observed or predicted with high/medium confidence level and having at least two cleavage sites associations were considered.

### Statistical analysis and definition of peptide markers

Statistical analyses were carried out using the statistical programming language R and the statistical software MedCalc version 12.7.5.0 (MedCalc Software; Mariakerke, Belgium). Signal intensities of each of the 3555 annotated peptides were correlated with serological laboratory markers using spearman’s rank correlation for univariate analysis. To reduce the risk of false discovery, the patient cohorts from Strasbourg and Paris were studied independently from each other. Statistical adjustment of p-values for multiple testing was performed by the method of Benjamini and Hochberg (24) and by taking into account the zero-inflated distribution of the peptide’s intensity values (24,25).

For the selection of age- and gender-matched normal individuals as well as age-, gender- and eGFR-matched CKD patients as reference groups to compare signal intensities of selected peptides, the R-script “MatchIt” (26) was used, which conducts nearest neighbor matching with logistic regression to estimate the propensity score. These reference human urinary peptide profiles were from normal individuals and CKD patients of previously described study populations recorded by CE-MS under identical operating conditions and instrument settings (27–29).

## Results

### Urinary peptides with association to CTX, BAP and osteocalcin

In a first step it was investigated if urinary peptides associated with bone metabolism can be detected in the setting of kidney transplantation. For this purpose, we analyzed 85 kidney transplant recipients from transplantation centers in Strasbourg and Paris for their individual peptide profiles by CE-MS. Demographic and clinical data for the two patient groups from Strasbourg (N=27) and Paris (n=58) are presented in table 1. As indicated by this table, patients in both groups are comparable in respect to clinical and demographic characteristics. For most of the patients, samples were taken within the first two years after transplantation. As the only differences it was found that steroid treatment is more prevalent in the Paris group and serum PTH is higher in the Strasbourg group. Patient characteristics are representative for recent transplant recipients.

The 85 individual peptide profiles were annotated to a reference list of 3,555 urinary peptides of high abundance in kidney transplantation patients. The signal intensities of these annotated peptides in each individual peptide profile were then correlated to the serum levels of the bone metabolism laboratory markers Beta-CrossLaps (CTX), osteocalcin and bone alkaline phosphatase (BAP) by Spearman rank sum analysis. As shown in the volcano plot in figure 1A, in the case of the Strasbourg cohort of 27 patients this resulted in the identification of 92 peptides with significant correlation (FDR < 0.05) to CTX of which 87 (95%) showed positive correlation with Spearman rho coefficients in the range of 0.58 to 0.82 and only five (5%) showed inverse regulation with Spearman rho coefficients in the range of -0.59 to -0.69. As shown in the Venn diagram of figure 1 B, 43 (47%) of these 92 peptides were also significantly correlated with BAP serum levels as second bone metabolism marker. Of note, all 43 peptides were positively correlated to both CTX and BAP. In the case of the Paris group of 58 patients, 590 urinary peptides are significantly associated with CTX. A volcano plot of Spearman rho values for urinary peptides of the Paris group is presented in figure 1 C. From these, 387 (66%) showed a positive correlation (range of Spearman rho values: 0.34 to 0.75), whereas 203 (34%) were found inversely regulated (range of Spearman rho values: -0.33 to -0.74). In the case of the Paris group, osteocalcin was available as a second bone marker. As shown in the Venn diagram of figure 1 D, an overlap of 455 (77%) peptides was observed showing to both CTX and osteocalcin a significant correlation in the same direction. Out of these, 293 demonstrated positive and 162 inverse correlation to both BR serum markers.

**Figure 1A.**
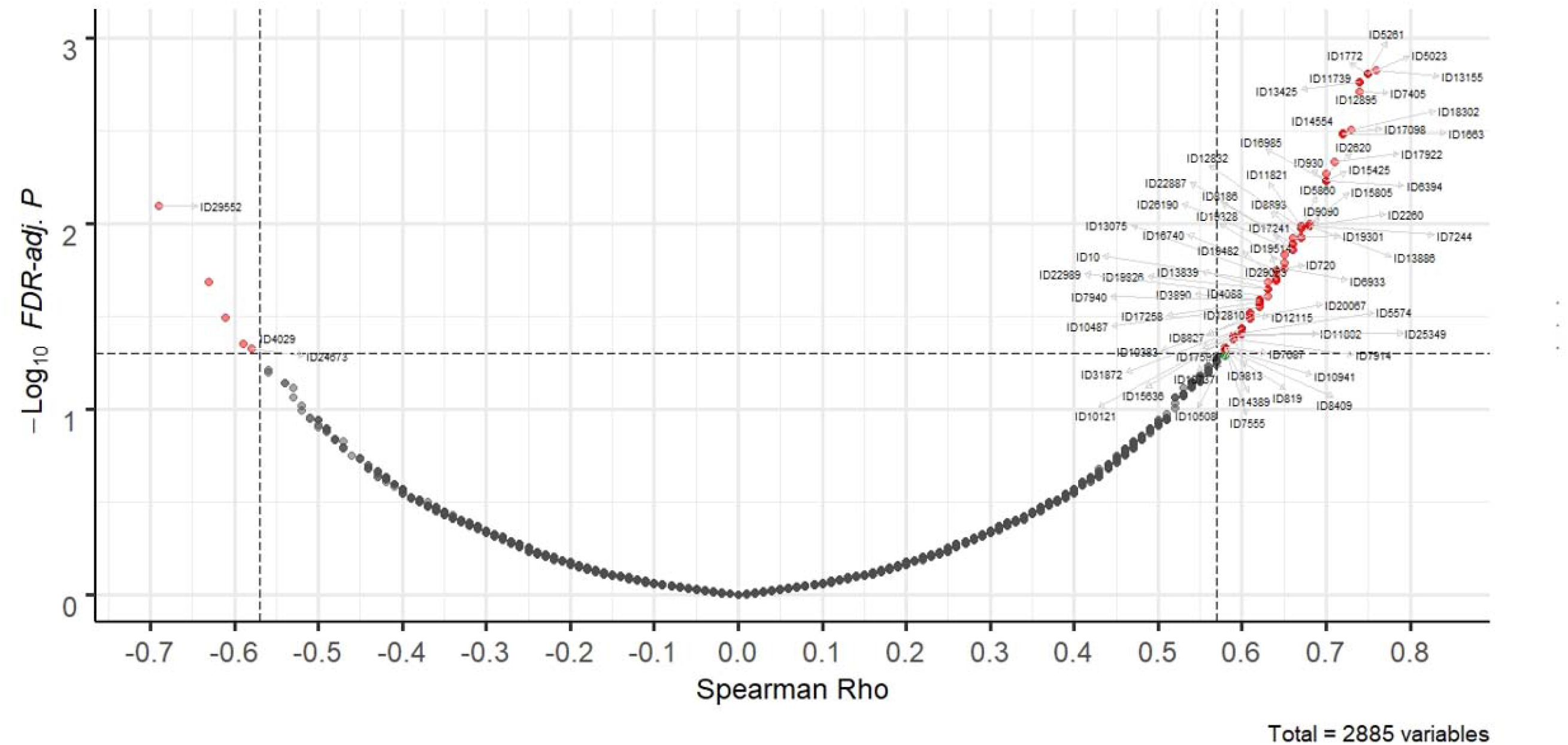
Volcano plot of Spearman Rho correlation factors for signal intensities of urinary peptides to CTX serum levels in the Strasbourg group of KTR patients. The 82 urinary peptides found in both the Strasbourg and Paris patient groups to be significantly correlated with CTX levels are labelled by their ID.

**Figure 1B.**
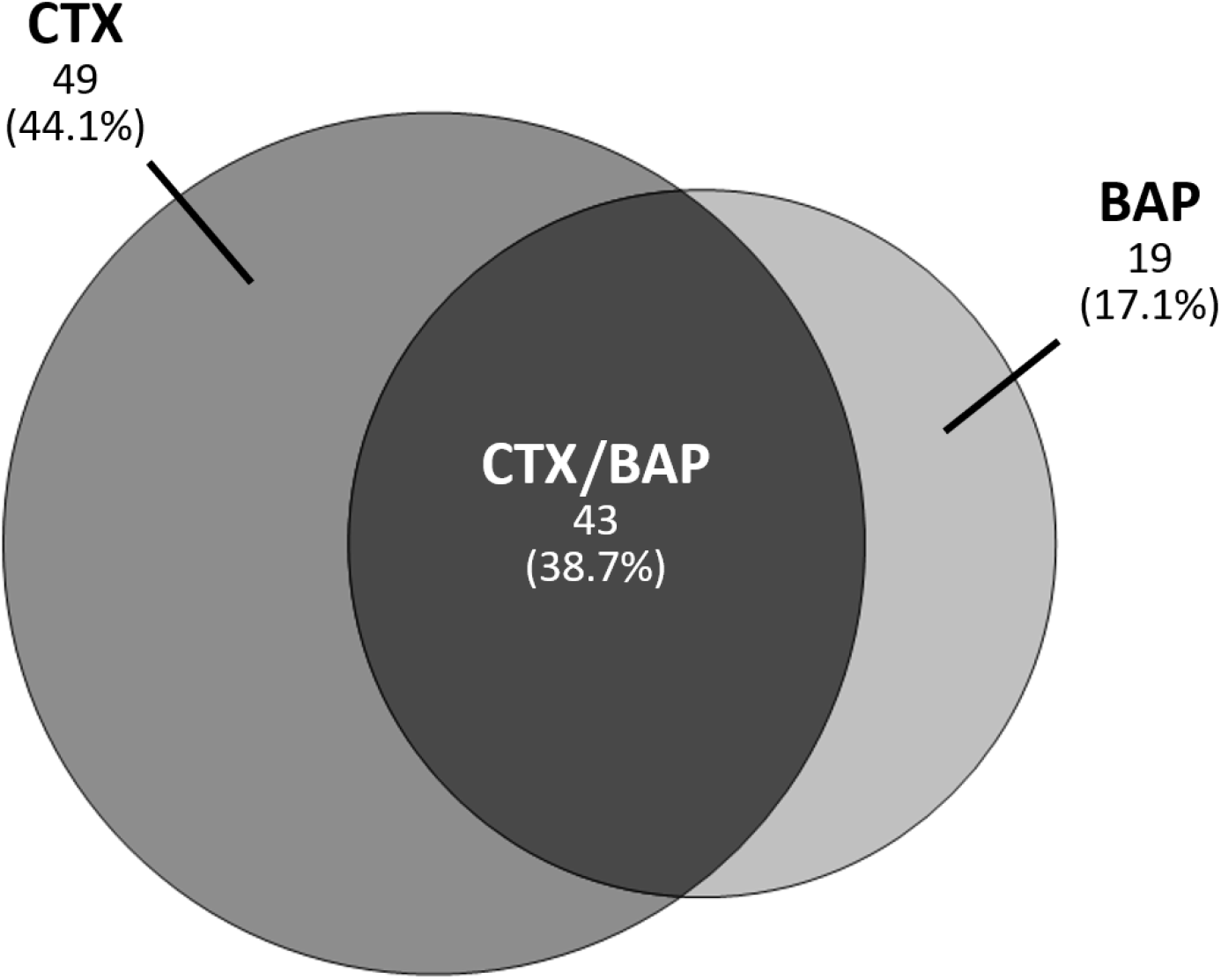
Venn diagram for urinary peptides with significant correlations to serum levels of Beta-CrossLaps (CTX) and bone alkaline phosphatase (BAP) in the Strasbourg group of KTR patients. In the Strasbourg KTR group, 92 peptides were identified for CTX and 62 for BAP out of which 43 were correlated to both serum markers for bone resorption. As revealed by the size of the circles, there is a higher prevalence of urinary peptides for correlation to the bone resorption marker CTX than to the bone formation marker BAP.

**Figure 1C.**
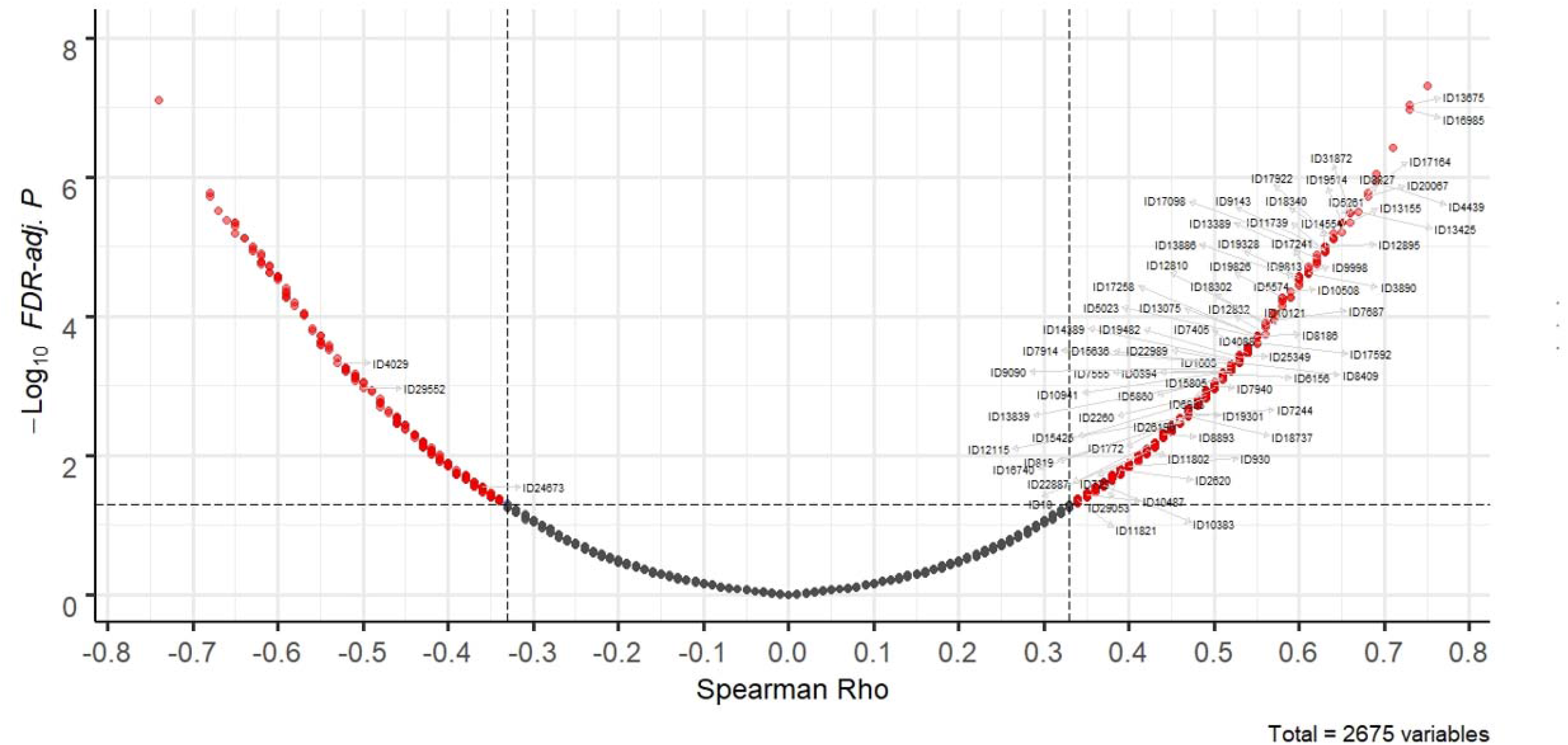
Volcano plot of Spearman Rho correlation factors for signal intensities of urinary peptides to CTX serum levels in the Paris group of KTR patients. The 82 urinary peptides found in both the Strasbourg and Paris patient groups to be significantly correlated with CTX levels are labelled by their ID.

**Figure 1D.**
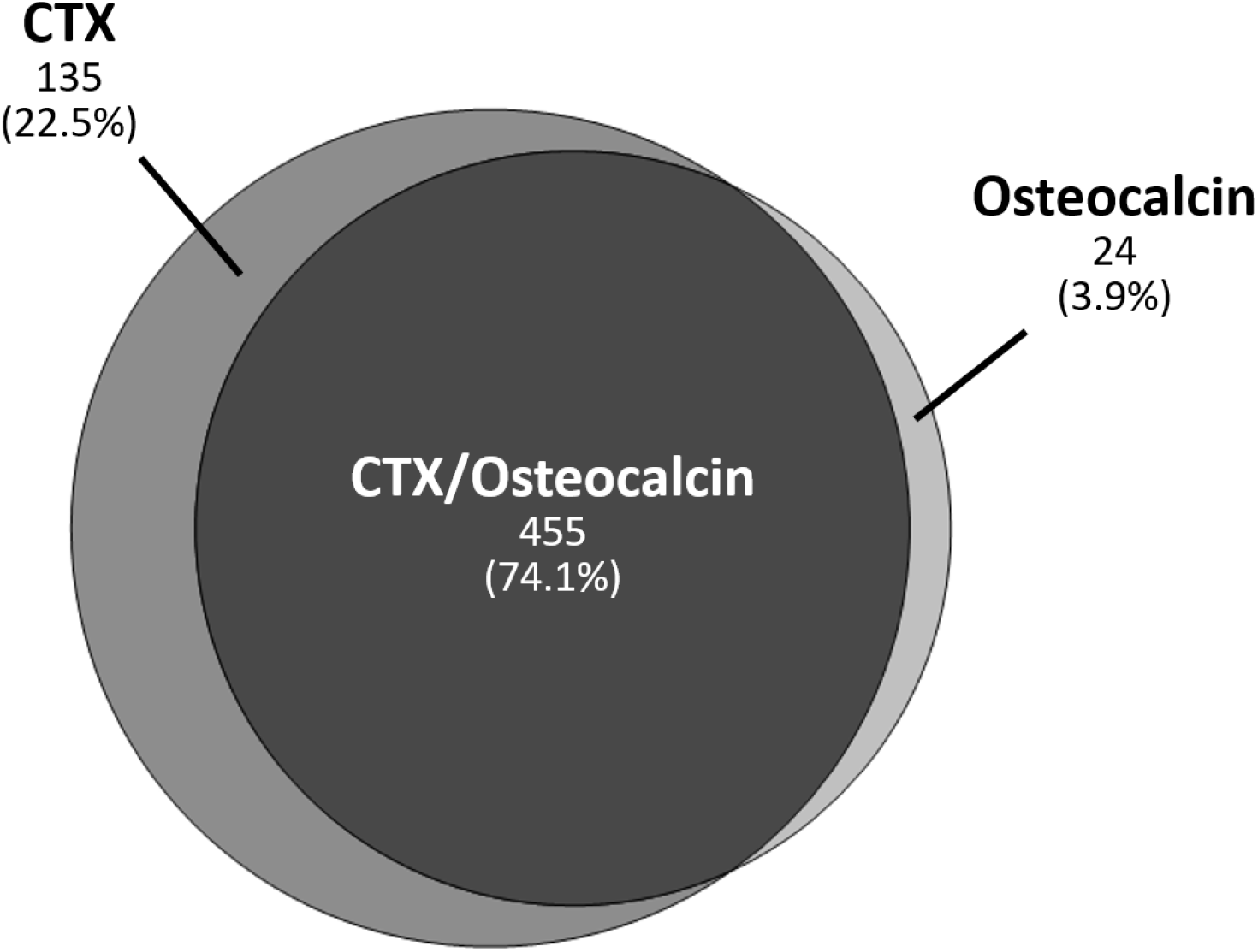
Venn diagram for urinary peptides with significant correlations to serum levels of Beta-CrossLaps (CTX) and osteocalcin in the Paris group of KTR patients. In the Paris KTR group, 590 peptides were identified for CTX and 479 for osteocalcin out of which 455 were correlated to both serum markers for bone remodeling. More prominent than in the case of the Strasbourg KTR, there is a higher proportion of urinary peptides correlated to CTX than to osteocalcin in the Paris KTR group, but with a greater overlap between the two serum bone resorption markers.

Eighty-two out of the pool of BR-associated urinary peptide candidates were found to be significantly correlated with CTX in both the Strasbourg and Paris patient groups, which represents 89 % (82 out of 92) of the total number of CTX-correlated peptides in the Strasbourg group. The 82 peptides are presented in supplementary table 1 by their CE-MS characteristics and group-specific signal distributions and are also labelled with their respective peptide ID’s in the volcano plots of figure 1 A for the patients from Paris and in figure 1 B for the patients from Strasbourg. As presented in figure 2 A, congruence between the Spearman Rho correlation factors of CTX and osteocalcin was observed for the Paris cohort. The same accounts for the Spearman Rho correlation factors of CTX and BAP in the Strasbourg cohort (Figure 2 B). Together this indicates that the 82 selected peptides are not only correlated to CTX, but also to other bone resorption markers. Conversely, none of these peptides were correlated with serum PTH and creatinine levels, as well as with the estimated glomerular filtration rate, neither in the Strasbourg nor in the Paris cohort.

**Figure 2A.**
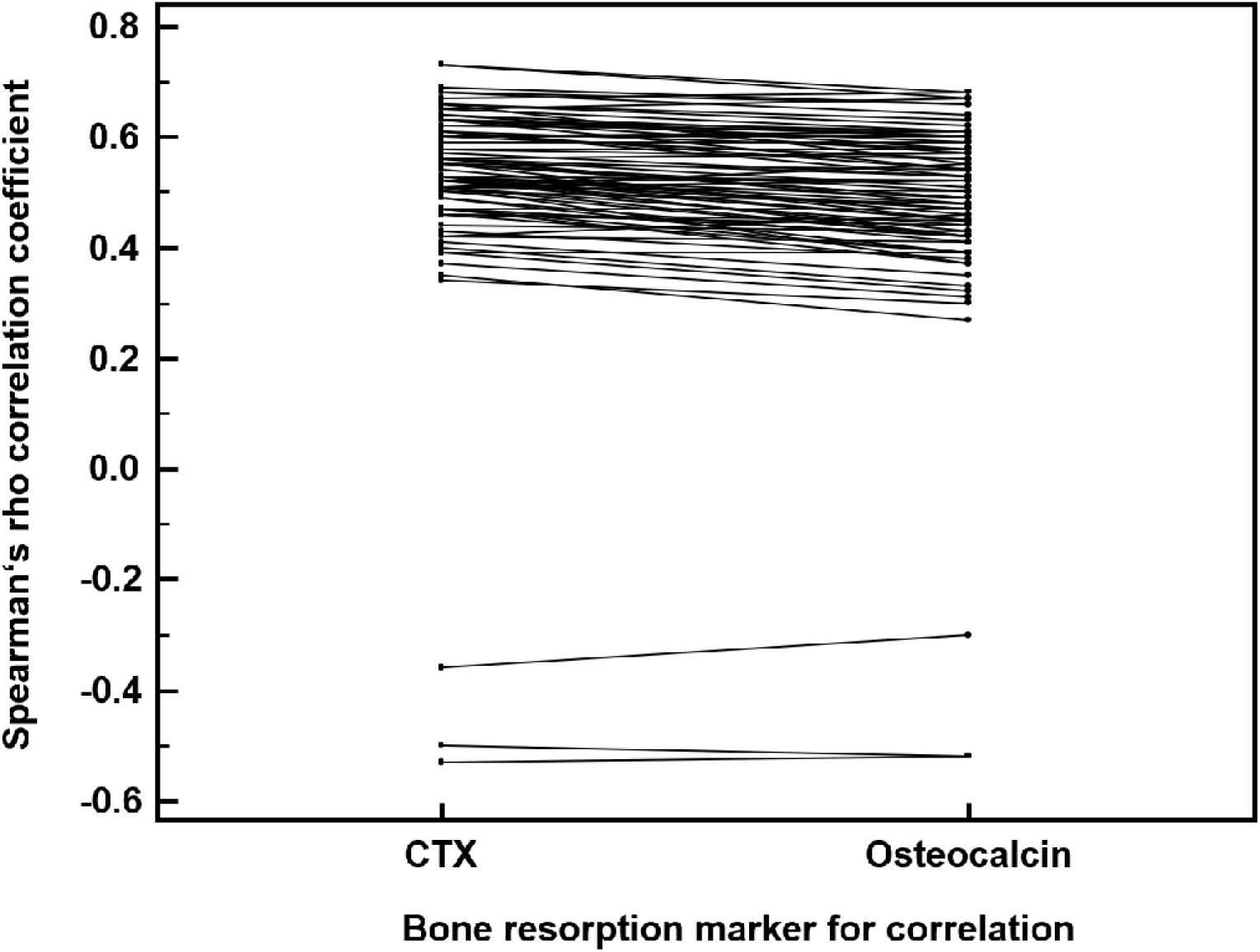
Pairwise comparison of Spearman Rho correlation factors to the serum bone remodeling markers CTX and osteocalcin for the selected 82 urinary peptides in the Paris KTR cohort. As revealed by this Wilcoxon signed rank test, all 82 urinary peptides selected due to their strong correlation to CTX, also show congruent correlation to osteocalcin.

**Figure 2B.**
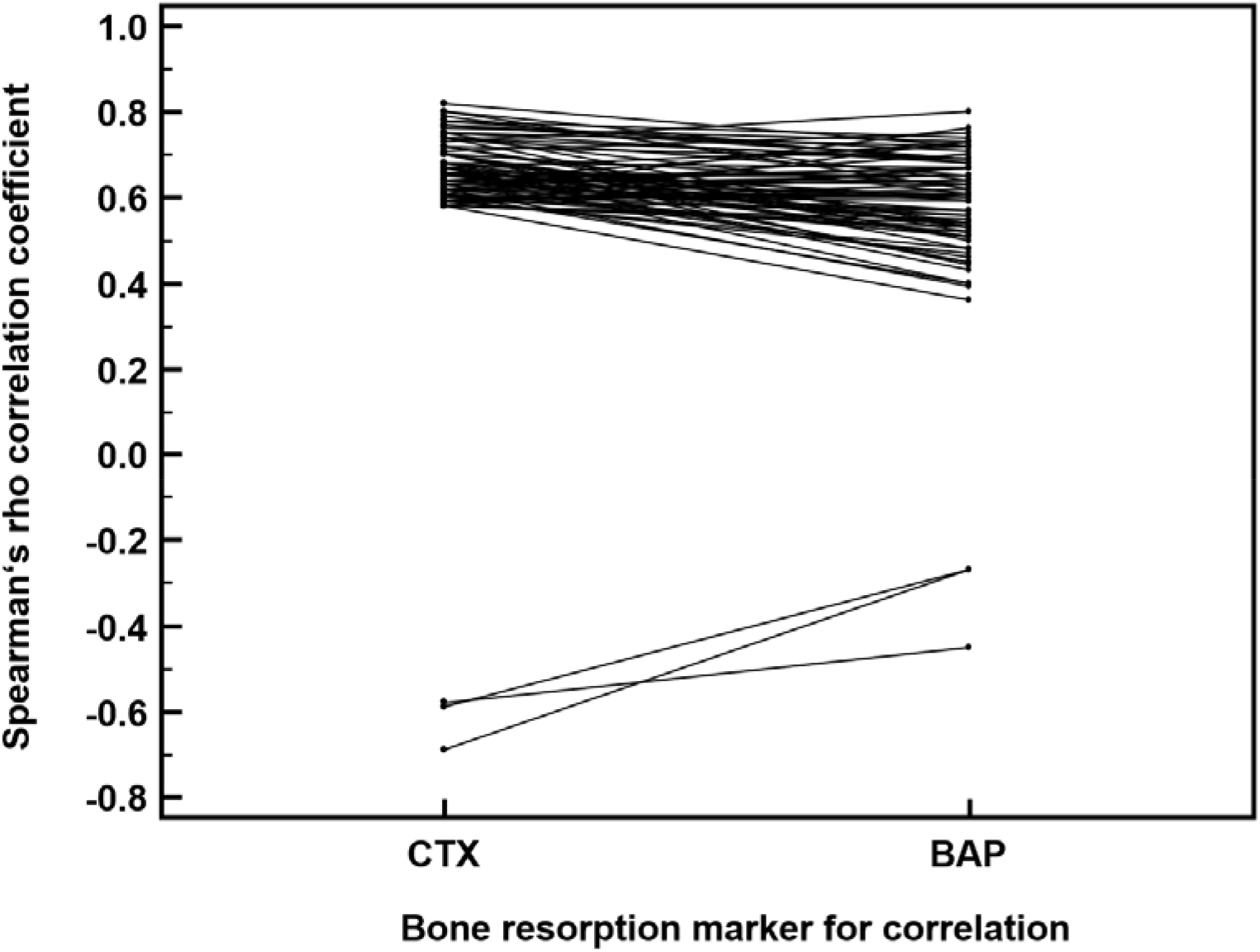
Pairwise comparison of Spearman Rho correlation factors to the serum bone resorption marker CTX and formation marker bone alkaline phosphatase (BAP) for the selected 82 urinary peptides in the Strasbourg KTR cohort. As revealed by this Wilcoxon signed rank test, all 82 urinary peptides selected due to their strong correlation to CTX, also show congruent correlation to BAP.

### CTX-associated urinary peptide identification and relation to bone metabolism

From these 82 CTX-associated urinary peptides, 54 (66%) could be resolved by their amino acid sequence. For these peptides sequence information is included in supplementary table 1. As can be deduced from this table, most of the CTX-associated peptides, namely 45, originate from alpha-1 type I collagen (COL1A1). From the remaining nine sequenced peptides, seven were derived from other collagen chains (COL1A2, n=1; COL2A1, n=1; COL3A1, n=2; COL4A2, n=1; COL4A5, n=2) and two from the non-collagen proteins EMID1 and gelsolin. All COL1A1 peptides belong to the group of peptides with positive correlation, whereas the COL3A1 peptides are two out of three peptides that are inversely correlated.

With 54.9% compared to 45.2%, the 45 CTX-associated COL1A1 urinary peptides contain to a significantly higher proportion hydroxyl modifications at proline residues than the 594 COL1A1 peptides in the total list of 3,555 peptides that are not correlated to CTX in both patient groups (p<0.0003 in a χ^2^-test). Moreover, as shown in figure 3, if the 45 peptide fragments are aligned to the linear sequence of COL1A1, it becomes evident that the peptides originate from nine hot spot regions. Since this observation can be interpreted as sign for regions of increased susceptibility to proteolytical cleavage, a search was carried out for observed and in silico predicted endoproteinase-specific cleavage sites in the immediate vicinity of these hot spots. By this kind of analysis, octapeptide sites matching with the consensus recognition sites of the proteases Cathepsin K (CTSK) and metalloproteinase 9 (MMP9) were identified by in silico-prediction using the software tool Proteasix, as indicated in figure 3. Further evidence for an involvement of CTSK in the generation of the BR-associated urinary peptides is provided by the finding that for the gelsolin-derived peptide 605-WVGTGASEAEKTGAQEL-621, as another BR-associated urinary peptide, cleavage by CTSK between leucin in position 604 and tryptophane in position 605 is reported by Vizovišek et al. (30).

**Figure 3.**
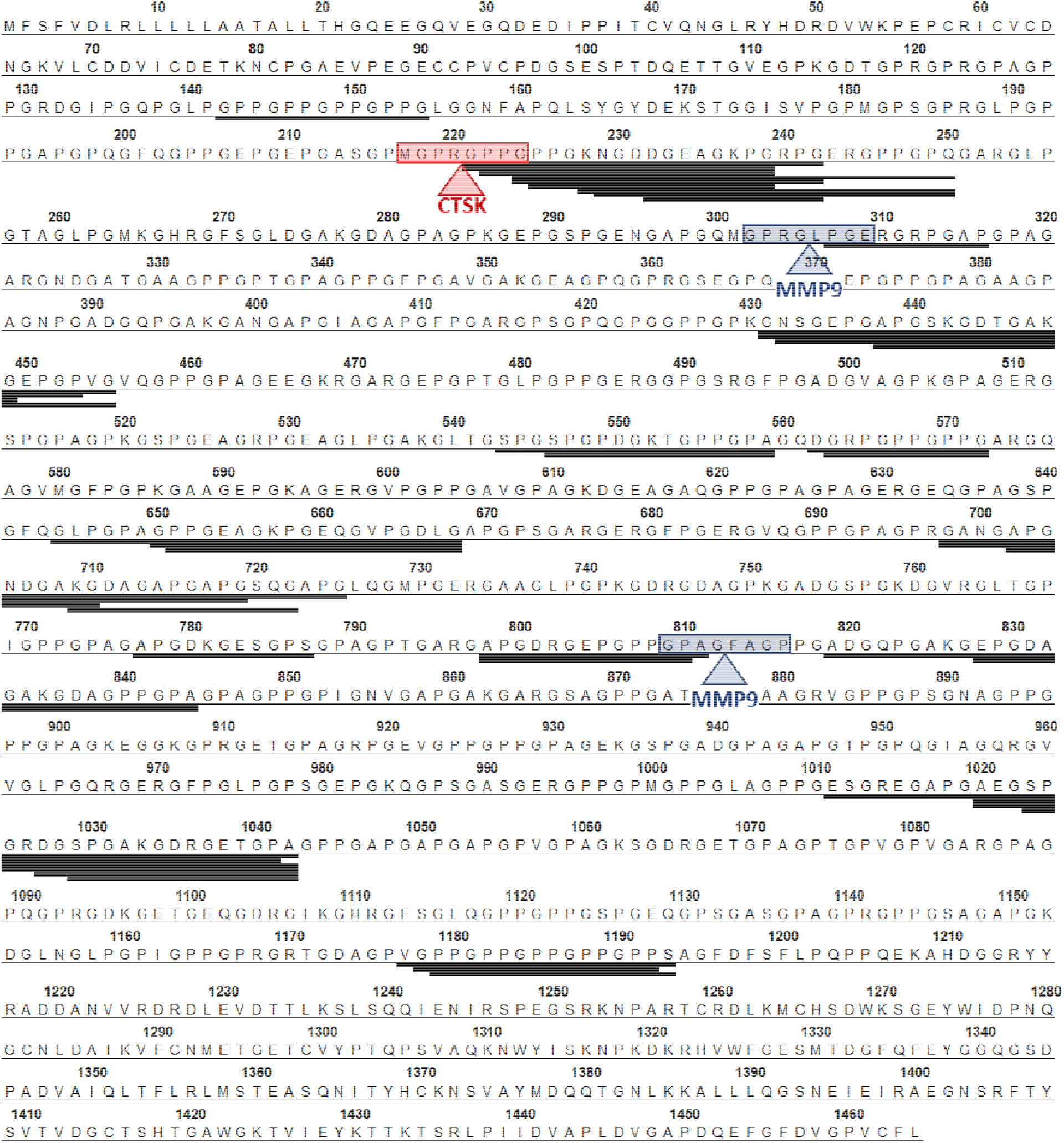
Alignment of the 45 COL1A1-derived peptides out of the 82 CTX-associated urinary peptides to the COL1A1 linear sequence. Cleavage sites for cathepsin K (CTSK) and matrix metalloproteinase-9 (MMP9) were discovered by in silico-prediction and are indicated by an arrow and by highlighting the octapeptide site matching with the consensus sequences for these proteases.

### Effect of bisphosphonate therapy on the CTX-associated urinary peptides

In order to evaluate sensitivity of the 82 CTX-associated urinary peptides to therapeutic interventions against bone loss, the distribution of these peptides before and after bisphosphonate treatment were determined in a separate group of 11 KTR enrolled by the university of Strasbourg. For bisphosphonate therapy, all but one patient received 70 mg oral alendronic acid once a week, whereas the remaining patient received 35 mg oral risedronate once a week. From the set of 82 peptides, 17 were sensitive to bisphosphonate treatment, all of which showed a decrease in their abundance after therapy (Table 2).

**Table 2.** Mean signal amplitudes and frequency of occurrence for the 17 bisphosphonate-sensitive peptides out of the set of 82 CTX-associated urinary peptides in 11 KTR patients before and after bisphosphonate (BP) treatment. All 17 peptides demonstrate reduced abundance after BP therapy. P-values were calculated based on pairwise comparison using the Wilcoxon signed rank test.

| Peptide ID<sup>‡</sup> | KTR Strasbourg Bisphosphonate (BP) treatment group (N=11) |  |  |  |  |
| --- | --- | --- | --- | --- | --- |
|  | Before BP therapy |  | After BP therapy |  | Two-tailed probability p<sup>†</sup> |
|  | Mean Amp (SD) | Freq | Mean Amp (SD) | Freq |  |
| 4088 | 1866 (3303) | 82 | 262 (841) | 18 | 0.0039 |
| 5261 | 16304 (7925) | 100 | 4988 (4170) | 91 | 0.001 |
| 5574 | 2814 (1655) | 91 | 593 (985) | 45 | 0.0049 |
| 7405 | 21806 (8966) | 100 | 6010 (6070) | 82 | 0.001 |
| 10508 | 23763 (11942) | 100 | 8063 (7816) | 82 | 0.001 |
| 13155 | 844 (1041) | 91 | 70 (186) | 18 | 0.0068 |
| 13675 | 2802 (1900) | 100 | 363 (598) | 36 | 0.001 |
| 13886 | 3040 (1614) | 91 | 906 (1353) | 55 | 0.002 |
| 14554 | 1044 (571) | 91 | 198 (383) | 27 | 0.0059 |
| 17098 | 1386 (899) | 91 | 152 (284) | 55 | 0.002 |
| 17164 | 2793 (2347) | 91 | 171 (348) | 36 | 0.002 |
| 18340 | 631 (772) | 73 | 79 (146) | 27 | 0.037 |
| 19328 | 7267 (5483) | 100 | 1057 (2591) | 64 | 0.001 |
| 19514 | 8890 (5213) | 100 | 1546 (3044) | 82 | 0.001 |
| 20067 | 2352 (1259) | 100 | 458 (733) | 45 | 0.001 |
| 22989 | 808 (820) | 73 | 162 (453) | 27 | 0.0078 |
| 25349 | 1119 (589) | 100 | 106 (239) | 27 | 0.001 |

**Abbreviations:** BP, Bisphosphonate; Freq, frequency of occurrence (%); KTR, kidney transplant recipient; Mean Amp, mean signal amplitude (ion counts); N, number of patients; SD, standard deviation.
‡ Peptide identification number.
† acc. Wilcoxon signed rank test

We compared the excretion levels of the 17 bisphosphonate-sensitive peptides of our KTR patient groups with those found in age-and gender-matched normal individuals, as well as age-, gender- and eGFR-matched chronic kidney disease patients. Since we were unable to match normal individuals also for eGFR, we included CKD patients into this comparative analysis in order to investigate the impact of eGFR on the peptide’s excretion levels. As presented in figure 4, overall excretion of these 17 peptides is significantly higher in KTR patients not treated with bisphosphonate compared to normal individuals but also CKD patients. After bisphosphonate therapy the direction of regulation reverts into the opposite, now being significantly lower in bisphosphonate treated KTR patients compared to both the normal individual and CKD patient reference groups.

**Figure 4.**
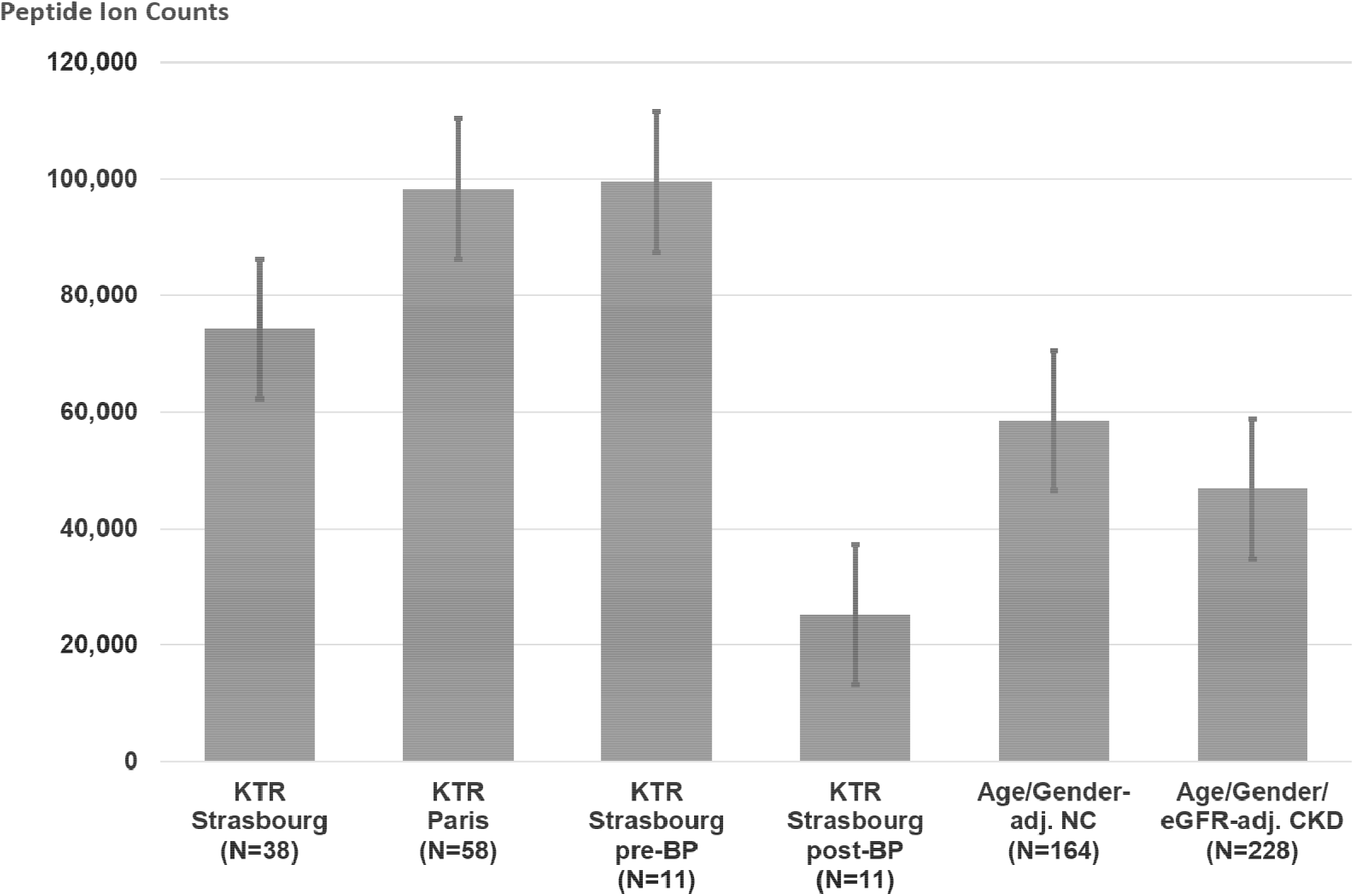
Overall summative excretion of the 17 bisphosphonate (BP)-sensitive urinary peptides in the different KTR study cohorts in comparison to age-and gender-matched normal controls (NC) and age-, gender-and eGFR-matched chronic kidney disease (CKD) patients.

## Discussion

Chronic kidney disease (CKD) is associated with mineral bone disease (CKD-MBD), a grouping of conditions including varying degrees of osteodystrophy, osteomalacia, adynamic bone, and abnormal bone mineralization(31,32). While several metabolic disorders improve after renal transplantation, bone damage persists and may even worsen(33). The mechanisms of this post-transplantation impairment are numerous and complex. The persistence of hyperparathyroidism and the initiation of anti-rejection treatment, mostly including corticosteroids, are important causes. Bone dynamics are here marked by an imbalance between the activity of bone resorption and bone formation causing a reduction in bone mineral density (BMD). Its fall predominates in the first year after transplantation (34) and contributes to a threefold increase in fracture risk (35).

Markers of bone remodeling are used to monitor patients at risk of osteoporosis and include markers of bone formation and bone resorption and both categories are further subdivided into non-collagen derived and collagen-derived markers. Their use has been integrated in the clinical dashboard of many transplantation departments, including ours (36,37). Serum levels of the C-terminal crosslinked telopeptide of collagen I (CTX), a resorption marker, have been shown to be associated with fracture risk in numerous studies (38–41).

In the present study we screened the urinary peptidome of KTRs looking for peptides correlated with CTX using conservative statistics and found a set of 82 peptides meeting the criteria in two independent groups of KTRs. By sequence analysis we found that most of these peptides are fragments of COL1A1, the most abundant protein in bone(42). It is therefore highly probable that CTX is not the only COL1A1 fragment that is released into the circulation as a result of bone metabolism. Whether they are freely filtered by the glomerulus, actively secreted by the tubule or released into urine by exosomal transport remains to be elucidated.

While all the COL1A1 peptides were positively correlated to CTX, two out of the three peptides showing negative correlation to CTX were fragments of COL3A1. Aside from collagen derived peptides, there were only two peptides derived from non-collagenous proteins. One is a fragment of gelsolin and the other of EMI domain-containing protein 1 (EMID1). Gelsolin is involved in the podosome assembly, a process used by several cell types including osteoclasts for matrix attachment and cytoskeletal reshaping (43). EMID1 has been shown to be an osteoblast marker in bone-forming metastases of prostate cancer(44). The peptide itself is derived from the collagen-like domain of EMID1 and has striking sequence similarities with other collagen peptides of our set, including 3 GPP amino acid sequence repeats.

Our results also shed light on the pathophysiology of bone remodeling. We found that the urinary CTX-associated COL1A1 peptides contain HyP in a significantly higher frequency than COL1A1 peptides not correlated to CTX. In addition, the analysis of the positions of HyP in the primary sequence of COL1A1 was in disagreement with that recorded in the public UNIPROT/SWISPROT database(45). This specific database entry relies on sequence similarity with chick skin collagen sequencing data from the 1970’s, not experimental data from human tissues (https://www.uniprot.org/uniprot/P02452). We found out that the positions of HyP in COL1A1 actually seem to be variable. We confirmed the variable position of COL1A1 HyP in other urine samples analyzed on different instruments in a different lab (personal data available on request). Others have recently reported similar findings in collagen I from other tissues(46). Explaining why CTX-associated COL1A1 peptides are more hydroxylated will certainly require understanding the causes underlying this variability in HyP positions. It has been shown that proline hydroxylation can take place in a non-enzymatic way due to reactive oxygen species, occurring as a *cis-*isomer instead of the enzymatically produced *trans-*isomer(47). This phenomenon could underlie our observation. Another hypothesis would be that there is a so far unknown, persistent prolyl-hydroxylase activity affecting collagen fibrils in the bone matrix. In any case, the link between collagen hydroxylation and bone resorption requires further investigation.

In addition, we found that most of the CTX-associated COL1A1 peptides originate from cluster regions, which hints towards the involvement of endoproteinases. Therefore, we searched databases of protease cleavage sites and we discovered that the most frequent cleavage signatures belong to Cathepsin K and MMP9. Cathepsin K is a cysteine protease and plays a pivotal role in osteoclast-mediated bone resorption (48). It degrades collagen I in the organic matrix of bone (49) and has become a drug target in the treatment of osteoporosis (50). Collagen I is also a known substrate of MMP9 a protease required for bone repair following fractures and involved in bone resorption in combination with other matrix metalloproteinases (51,52). Altogether these results strengthen the hypothesis that the selected peptides were generated through osteoclast-mediated bone resorption.

We then tested the effect of bisphosphonates on the urinary peptides of interest. Bisphosphonates are stable analogues of inorganic pyrophosphate that exhibit strong affinity for the bone mineral matrix and inhibit the functioning of osteoclasts (53). This process results in an overall slowing of bone turnover and prevents bone loss, thereby reducing the incidence of osteoporotic fractures. Because we had strong arguments in favor of an osseous origin of the urinary CTX-associated peptides, we tried to replicate the results of a seminal experiment from 1971 showing the effect of bisphosphonates on urinary output of HyP-containing peptides (54,55). In that study sodium etidronate was administered orally to four patients with Paget’s disease. As a result, previously elevated serum levels of alkaline phosphatase and both plasma and urinary HyP decreased by more than half after three months of treatment. Urinary HyP was present in the form of peptides whose sequence remained unknown. These results suggested that the reduction in bone resorption following treatment with bisphosphonates was accompanied by a decrease in collagen fragments (possibly derived from resorption) in the blood and urine. According to a similar study scheme, we compared the urinary levels of the 82 CTX-associated peptides before and after treatment with oral bisphosphonates administered during several months to KTRs at risk of fractures. As we expected, the abundance of 14 HyP-rich type I collagen fragments (and 3 peptides of unknown sequence) significantly and sharply declined (on average by 82 %) following bisphosphonate treatment.

Our study has limitations. The association between CTX and urinary peptides was different in the two KTR cohorts, with a far greater number of peptides correlating with bone data in the Paris cohort than in the Strasbourg cohort (see volcano plots, figure 1A and 1C), partly due to cohort size and subsequent statistical power differences. This reduced the number of peptides in our peptide set but suggests that there could be far more peptides associated with BR than we report. Moreover, the bisphosphonate study was small with only 11 patients. However, the decrease in the urinary levels of 17 peptides was so dramatic following bisphosphonate treatment that it partly overcame the problem of this limited statistical power. Here too, we hypothesize that more bisphosphonate-sensitive peptides can be identified by increasing the number of study subjects.

Our observations combined with established knowledge on bone remodeling strongly suggest that peptides whose urinary abundance strongly correlates with serum CTX levels originate through bisphosphonate-sensitive bone mechanisms, possibly bone resorption. This hypothesis is in agreement with results from older studies using isotopes to trace the fate of collagen metabolism and showing that HyP-containing peptides from bone are released into the blood stream following bone resorption and finally end up in urine (56–59). The use of these peptides as biomarkers could become complementary to that of existing bone remodeling markers in this KTR population.

The key question for clinicians will be whether these urinary peptides predict the evolution of bone density and occurrence of fractures in this high-risk population. Finally, our study focuses on KTRs but whether the results we observed in this population can be extrapolated to other populations remains to be determined in future studies.

## Data Availability

All data produced in the present study are available upon reasonable request to the authors

## Notes

### Competing Interest Statement

The authors have declared no competing interest.

### Funding Statement

This study did not receive any funding

### Author Declarations

The study was approved by the institutional review board of Strasbourg University Hospitals under the reference CPP-EST DC-2013-1990 for patients from Strasbourg. For patients from the Necker Hospital the study was approved by the Ethics Committee of Ile-de-France XI (#13016).

