## Supplemental Table 1 for "Urinary collagen-derived peptides as sensitive markers for bone resorption and bisphosphonate treatment in kidney transplant patients"

**Supplementary Table 2.** Characteristics of CTX-associated urinary peptides identified in KTR groups of the Paris Necker and Strasbourg university hospitals by capillary electrophoresis coupled to mass spectrometry.

| **CE-MS characteristics** | | | | |  | **Peptide distribution** | | | | | | |  | **Sequence information** | | | | | | |
| --- | --- | --- | --- | --- | --- | --- | --- | --- | --- | --- | --- | --- | --- | --- | --- | --- | --- | --- | --- | --- |
| **Peptide ID^‡^** |  | **Molecular mass (Dalton)** |  | **CE-migration time (minutes)** |  | **KTR Paris**  **(N=58)** | | |  | **KTR Strasbourg**  **(N=38)** | | |  | **Protein**  **sequence^†^** |  | **Protein**  **Symbol^§^** |  | **AA^***^** |  | **Theoretical Mass** |
|  |  |  |  |  |  | **Mean Amp (SD)** |  | **Freq** |  | **Mean Amp**  **(SD)** |  | **Freq** |  |  |  |  |  |  |  |  |
| 10 |  | 801.354 |  | 23.16 |  | 15 (30) |  | 28 |  | 91 (182) |  | 59 |  | ApGNDGAKG |  | COL1A1 |  | 702 - 710 |  | 801.361 |
| 720 |  | 924.430 |  | 33.98 |  | 31 (130) |  | 16 |  | 44 (75) |  | 37 |  | --- |  | --- |  | --- |  | --- |
| 819 |  | 935.438 |  | 23.96 |  | 183 (283) |  | 64 |  | 439 (679) |  | 78 |  | GRpGPpGPpG |  | COL1A1 |  | 563 - 572 |  | 935.446 |
| 930 |  | 945.401 |  | 25.76 |  | 202 (218) |  | 78 |  | 193 (271) |  | 74 |  | --- |  | --- |  | --- |  | --- |
| 1663 |  | 1008.505 |  | 23.03 |  | 35 (58) |  | 38 |  | 33 (62) |  | 37 |  | pGERGRPGAP |  | COL1A1 |  | 307 - 316 |  | 1008.510 |
| 1772 |  | 1016.440 |  | 25.77 |  | 2863 (2554) |  | 97 |  | 4264 (5581) |  | 85 |  | ApGDKGESGPS |  | COL1A1 |  | 777 - 787 |  | 1016.441 |
| 2260 |  | 1050.471 |  | 26.95 |  | 708 (801) |  | 90 |  | 855 (1231) |  | 89 |  | DGRpGPpGPpG |  | COL1A1 |  | 562 - 572 |  | 1050.473 |
| 2620 |  | 1071.493 |  | 21.41 |  | 184 (203) |  | 67 |  | 103 (146) |  | 52 |  | DGEAGKpGRpG |  | COL1A1 |  | 232 - 242 |  | 1071.495 |
| 3890 |  | 1134.584 |  | 23.96 |  | 629 (560) |  | 90 |  | 523 (759) |  | 63 |  | PIGQEGAPGRPG |  | COL4A2 |  | 1463 - 1474 |  | 1134.578 |
| 4029 |  | 1140.478 |  | 21.17 |  | 2467 (2498) |  | 67 |  | 2048 (1491) |  | 78 |  | DKGDTGPPGPQG |  | COL3A1 |  | 628 - 639 |  | 1124.510 |
| 4088 |  | 1143.514 |  | 36.93 |  | 678 (1399) |  | 86 |  | 343 (298) |  | 85 |  | GpPGPpGPpGPpG |  | COL1A1 |  | 142 - 154 |  | 1143.520 |
| 4439 |  | 1162.542 |  | 20.25 |  | 398 (383) |  | 83 |  | 267 (268) |  | 74 |  | --- |  | --- |  | --- |  | --- |
| 5023 |  | 1188.529 |  | 24.20 |  | 92 (116) |  | 53 |  | 85 (116) |  | 56 |  | --- |  | --- |  | --- |  | --- |
| 5261 |  | 1200.538 |  | 24.50 |  | 17913 (11916) |  | 100 |  | 12317 (9098) |  | 96 |  | KGDAGApGApGSQG |  | COL1A1 |  | 709 - 722 |  | 1200.537 |
| 5574 |  | 1216.537 |  | 24.61 |  | 3420 (2103) |  | 100 |  | 2045 (1699) |  | 85 |  | --- |  | --- |  | --- |  | --- |
| 5860 |  | 1232.542 |  | 24.78 |  | 68 (80) |  | 53 |  | 52 (79) |  | 37 |  | --- |  | --- |  | --- |  | --- |
| 6156 |  | 1250.561 |  | 27.77 |  | 49218 (19867) |  | 100 |  | 38763 (21000) |  | 100 |  | ApGDRGEpGPPGp |  | COL1A1 |  | 798 - 810 |  | 1250.553 |
| 6394 |  | 1265.590 |  | 27.30 |  | 14293 (13217) |  | 97 |  | 15696 (23651) |  | 93 |  | SpGPDGKTGPpGPA |  | COL1A1 |  | 546 - 559 |  | 1265.589 |
| 6933 |  | 1296.594 |  | 19.33 |  | 412 (635) |  | 64 |  | 297 (346) |  | 67 |  | --- |  | --- |  | --- |  | --- |
| 7244 |  | 1314.613 |  | 21.76 |  | 638 (602) |  | 78 |  | 536 (600) |  | 82 |  | SpGAKGDRGETGPA |  | COL1A1 |  | 1029 - 1042 |  | 1314.616 |
| 7405 |  | 1321.595 |  | 28.25 |  | 25229 (16821) |  | 100 |  | 17013 (14806) |  | 96 |  | ApGDRGEpGPpGPA |  | COL1A1 |  | 798 - 811 |  | 1321.590 |
| 7555 |  | 1329.595 |  | 22.59 |  | 77 (148) |  | 36 |  | 19 (30) |  | 30 |  | --- |  | --- |  | --- |  | --- |
| 7687 |  | 1337.620 |  | 38.21 |  | 186 (454) |  | 53 |  | 55 (96) |  | 44 |  | GPpGPpGPpGpPGPP |  | COL1A1 |  | 1178 - 1192 |  | 1337.625 |
| 7914 |  | 1351.636 |  | 38.75 |  | 394 (532) |  | 83 |  | 1466 (3859) |  | 78 |  | PpGPpGPpGPPGPPS |  | COL1A1 |  | 1179 - 1193 |  | 1351.641 |
| 7940 |  | 1353.655 |  | 25.96 |  | 509 (400) |  | 91 |  | 377 (315) |  | 82 |  | PVGpSGKDGANGIpG |  | COL1A1 |  | 1168 - 1182 |  | 1353.652 |
| 8186 |  | 1367.642 |  | 38.82 |  | 2149 (2139) |  | 93 |  | 8227 (24698) |  | 93 |  | pPGpPGPpGpPGPPS |  | COL1A1 |  | 1179 - 1193 |  | 1367.636 |
| 8409 |  | 1383.636 |  | 38.97 |  | 256 (328) |  | 69 |  | 1372 (5559) |  | 48 |  | --- |  | --- |  | --- |  | --- |
| 8827 |  | 1407.665 |  | 37.25 |  | 388 (551) |  | 76 |  | 281 (364) |  | 59 |  | PGLpGFpGTpGpPGP |  | COL4A5 |  | 1118 - 1132 |  | 1407.667 |
| 8893 |  | 1410.650 |  | 22.70 |  | 42 (106) |  | 28 |  | 90 (160) |  | 33 |  | pPGKNGDDGEAGKPG |  | COL1A1 |  | 225 - 239 |  | 1410.637 |
| 9090 |  | 1423.652 |  | 37.40 |  | 232 (345) |  | 53 |  | 422 (1026) |  | 56 |  | GpSGVpGQpGSPGLpG |  | COL4A5 |  | 1045 - 1060 |  | 1423.658 |
| 9143 |  | 1426.640 |  | 22.66 |  | 245 (389) |  | 64 |  | 477 (940) |  | 74 |  | PpGKNGDDGEAGKpG |  | COL1A1 |  | 225 - 239 |  | 1426.632 |
| 9813 |  | 1470.684 |  | 21.18 |  | 152 (208) |  | 62 |  | 117 (266) |  | 48 |  | --- |  | --- |  | --- |  | --- |
| 9998 |  | 1486.683 |  | 21.27 |  | 626 (655) |  | 85 |  | 477 (567) |  | 74 |  | DGSpGAKGDRGETGPA |  | COL1A1 |  | 1027 - 1042 |  | 1486.665 |
| 10121 |  | 1494.661 |  | 30.22 |  | 462 (798) |  | 85 |  | 367 (336) |  | 78 |  | EpGDAGAKGDAGPpGPA |  | COL1A1 |  | 828 - 844 |  | 1494.659 |
| 10383 |  | 1513.639 |  | 29.74 |  | 175 (262) |  | 53 |  | 250 (658) |  | 52 |  | --- |  | --- |  | --- |  | --- |
| 10487 |  | 1522.705 |  | 29.53 |  | 1250 (801) |  | 93 |  | 1369 (1138) |  | 85 |  | SpGSpGPDGKTGPpGPA |  | COL1A1 |  | 543 - 559 |  | 1522.690 |
| 10508 |  | 1523.733 |  | 39.95 |  | 18652 (9778) |  | 100 |  | 14187 (6918) |  | 100 |  | VGPpGPpGPpGPpGPPS |  | COL1A1 |  | 1177 - 1193 |  | 1523.726 |
| 10941 |  | 1554.641 |  | 28.80 |  | 88 (101) |  | 57 |  | 59 (77) |  | 63 |  | --- |  | --- |  | --- |  | --- |
| 11739 |  | 1612.761 |  | 23.44 |  | 238 (368) |  | 53 |  | 264 (374) |  | 52 |  | ApGSKGDTGAKGEpGPVG |  | COL1A1 |  | 438 - 455 |  | 1612.769 |
| 11802 |  | 1617.721 |  | 23.26 |  | 83 (135) |  | 40 |  | 65 (100) |  | 37 |  | NSGEpGApGSKGDTGAKG |  | COL1A1 |  | 432 - 449 |  | 1617.723 |
| 11821 |  | 1619.650 |  | 36.47 |  | 42 (124) |  | 14 |  | 170 (278) |  | 37 |  | GPPGPPGPMGPPGPPGPTG |  | EMID1 |  | 296 - 314 |  | 1619.777 |
| 12115 |  | 1640.580 |  | 23.00 |  | 8368 (5109) |  | 100 |  | 5876 (5006) |  | 96 |  | --- |  | --- |  | --- |  | --- |
| 12810 |  | 1688.662 |  | 20.11 |  | 212 (434) |  | 41 |  | 96 (166) |  | 44 |  | --- |  | --- |  | --- |  | --- |
| 12832 |  | 1689.806 |  | 27.93 |  | 460 (1837) |  | 60 |  | 255 (377) |  | 59 |  | GpAGPpGEAGKPGEQGVPG |  | COL1A1 |  | 647 - 665 |  | 1689.796 |
| 12895 |  | 1693.765 |  | 23.47 |  | 447 (531) |  | 69 |  | 1530 (3950) |  | 67 |  | PpGPpGKNGDDGEAGKpG |  | COL1A1 |  | 222 - 239 |  | 1693.754 |
| 13075 |  | 1708.778 |  | 30.76 |  | 2473 (1465) |  | 98 |  | 1981 (1481) |  | 100 |  | pPGEAGKpGEQGVpGDLG |  | COL1A1 |  | 651 - 668 |  | 1708.790 |
| 13155 |  | 1716.770 |  | 28.30 |  | 797 (801) |  | 83 |  | 602 (600) |  | 78 |  | --- |  | --- |  | --- |  | --- |
| 13389 |  | 1732.767 |  | 28.35 |  | 4298 (2528) |  | 97 |  | 3519 (3040) |  | 85 |  | WVGTGASEAEKTGAQEL |  | GSN |  | 605 - 621 |  | 1732.827 |
| 13425 |  | 1734.792 |  | 23.65 |  | 606 (1517) |  | 52 |  | 234 (563) |  | 52 |  | GppGPPGKNGDDGEAGKPG |  | COL1A1 |  | 221 - 239 |  | 1734.781 |
| 13675 |  | 1750.786 |  | 23.88 |  | 3645 (3522) |  | 91 |  | 3452 (3924) |  | 82 |  | GpPGpPGKNGDDGEAGKpG |  | COL1A1 |  | 221 - 239 |  | 1750.776 |
| 13839 |  | 1761.692 |  | 19.39 |  | 527 (692) |  | 71 |  | 308 (415) |  | 67 |  | --- |  | --- |  | --- |  | --- |
| 13886 |  | 1765.806 |  | 30.96 |  | 3534 (2373) |  | 98 |  | 2722 (2605) |  | 74 |  | GpPGEAGKpGEQGVpGDLG |  | COL1A1 |  | 650 - 668 |  | 1765.812 |
| 14389 |  | 1809.830 |  | 20.99 |  | 477 (820) |  | 62 |  | 120 (248) |  | 41 |  | GPpGKNGDDGEAGKpGRpG |  | COL1A1 |  | 224 - 242 |  | 1809.824 |
| 14554 |  | 1825.788 |  | 31.81 |  | 900 (903) |  | 85 |  | 941 (916) |  | 78 |  | GANGApGNDGAKGDAGApGApG |  | COL1A1 |  | 698 - 719 |  | 1825.783 |
| 15425 |  | 1899.862 |  | 21.43 |  | 457 (556) |  | 67 |  | 313 (402) |  | 70 |  | SpGRDGSpGAKGDRGETGPA |  | COL1A1 |  | 1023 - 1042 |  | 1899.867 |
| 15636 |  | 1916.770 |  | 20.23 |  | 4938 (4626) |  | 95 |  | 3209 (3275) |  | 89 |  | --- |  | --- |  | --- |  | --- |
| 15805 |  | 1932.722 |  | 34.66 |  | 153 (263) |  | 48 |  | 282 (591) |  | 37 |  | --- |  | --- |  | --- |  | --- |
| 16740 |  | 2034.870 |  | 19.50 |  | 326 (774) |  | 31 |  | 104 (243) |  | 30 |  | --- |  | --- |  | --- |  | --- |
| 16985 |  | 2058.942 |  | 23.27 |  | 845 (1080) |  | 83 |  | 493 (588) |  | 59 |  | --- |  | --- |  | --- |  | --- |
| 17098 |  | 2070.914 |  | 25.45 |  | 1213 (892) |  | 95 |  | 1247 (933) |  | 89 |  | GNSGEpGApGSKGDTGAKGEPGp |  | COL1A1 |  | 431 - 453 |  | 2070.909 |
| 17164 |  | 2076.955 |  | 21.79 |  | 2964 (2834) |  | 85 |  | 2385 (2744) |  | 82 |  | GPpGPpGKNGDDGEAGKpGRpG |  | COL1A1 |  | 221 - 242 |  | 2076.946 |
| 17241 |  | 2085.931 |  | 22.05 |  | 10892 (10975) |  | 72 |  | 7377 (9223) |  | 44 |  | AEGSpGRDGSpGAKGDRGETGP |  | COL1A1 |  | 1020 - 1041 |  | 2085.931 |
| 17258 |  | 2087.849 |  | 19.44 |  | 1293 (2496) |  | 71 |  | 872 (1096) |  | 78 |  | --- |  | --- |  | --- |  | --- |
| 17592 |  | 2124.845 |  | 20.34 |  | 441 (697) |  | 60 |  | 333 (544) |  | 59 |  | --- |  | --- |  | --- |  | --- |
| 17922 |  | 2156.978 |  | 22.21 |  | 2837 (2577) |  | 93 |  | 1856 (1522) |  | 85 |  | AEGSpGRDGSpGAKGDRGETGPA |  | COL1A1 |  | 1020 - 1042 |  | 2156.968 |
| 18302 |  | 2191.994 |  | 22.34 |  | 957 (910) |  | 86 |  | 600 (632) |  | 74 |  | NGDDGEAGKpGRpGERGpPGPQ |  | COL1A1 |  | 229 - 250 |  | 2191.984 |
| 18340 |  | 2194.973 |  | 20.16 |  | 847 (1200) |  | 78 |  | 746 (939) |  | 78 |  | NDGPpGRDGQpGHKGERGYpG |  | COL1A2 |  | 932 - 952 |  | 2194.974 |
| 18737 |  | 2226.995 |  | 26.19 |  | 27077 (19278) |  | 83 |  | 20120 (19872) |  | 56 |  | GNSGEpGApGSKGDTGAKGEPGpVG |  | COL1A1 |  | 431 - 455 |  | 2226.999 |
| 19301 |  | 2274.048 |  | 33.60 |  | 211 (259) |  | 72 |  | 91 (215) |  | 33 |  | GLpGpAGpPGEAGKPGEQGVpGDLG |  | COL1A1 |  | 644 - 668 |  | 2274.076 |
| 19328 |  | 2276.015 |  | 27.19 |  | 5293 (5376) |  | 72 |  | 5524 (5132) |  | 85 |  | ADGQPGAKGEpGDAGAKGDAGPPGpA |  | COL1A1 |  | 819 - 844 |  | 2276.030 |
| 19482 |  | 2289.030 |  | 33.69 |  | 223 (293) |  | 67 |  | 185 (504) |  | 59 |  | --- |  | --- |  | --- |  | --- |
| 19514 |  | 2292.022 |  | 27.22 |  | 8348 (5707) |  | 100 |  | 7507 (6181) |  | 93 |  | ADGQpGAKGEPGDAGAKGDAGppGPA |  | COL1A1 |  | 819 - 844 |  | 2292.025 |
| 19826 |  | 2320.077 |  | 20.79 |  | 351 (544) |  | 55 |  | 182 (283) |  | 52 |  | KNGDDGEAGKpGRPGERGppGPQ |  | COL1A1 |  | 228 - 250 |  | 2320.079 |
| 20067 |  | 2339.008 |  | 33.97 |  | 1986 (1911) |  | 90 |  | 1676 (1887) |  | 85 |  | GANGApGNDGAKGDAGApGApGSQGApG |  | COL1A1 |  | 698 - 725 |  | 2339.001 |
| 22887 |  | 2633.088 |  | 34.42 |  | 19 (52) |  | 21 |  | 41 (55) |  | 48 |  | --- |  | --- |  | --- |  | --- |
| 22989 |  | 2644.219 |  | 21.18 |  | 1664 (1752) |  | 81 |  | 833 (971) |  | 70 |  | GPpGKNGDDGEAGKPGRpGERGPpGpQ |  | COL1A1 |  | 224 - 250 |  | 2644.222 |
| 24673 |  | 2825.271 |  | 24.33 |  | 18312 (11444) |  | 98 |  | 18662 (11210) |  | 96 |  | ERGEAGIpGVpGAKGEDGKDGSpGEpGANG |  | COL3A1 |  | 448 - 477 |  | 2825.270 |
| 25349 |  | 2912.174 |  | 25.48 |  | 1241 (821) |  | 95 |  | 779 (681) |  | 82 |  | --- |  | --- |  | --- |  | --- |
| 26190 |  | 3013.293 |  | 22.22 |  | 10177 (9447) |  | 79 |  | 7765 (7748) |  | 85 |  | ESGREGApGAEGSPGRDGSpGAKGDRGETGpA |  | COL1A1 |  | 1011 - 1042 |  | 3013.336 |
| 29053 |  | 3378.361 |  | 38.40 |  | 160 (338) |  | 43 |  | 434 (696) |  | 67 |  | --- |  | --- |  | --- |  | --- |
| 29552 |  | 3441.600 |  | 31.33 |  | 11882 (6280) |  | 97 |  | 10556 (5931) |  | 100 |  | --- |  | --- |  | --- |  | --- |
| 31872 |  | 3815.810 |  | 21.90 |  | 1188 (1675) |  | 59 |  | 1039 (1508) |  | 63 |  | --- |  | --- |  | --- |  | --- |
| **Abbreviations:** AA, amino acid; CE, capillary electrophoresis; Freq, frequency of occurrence (%); KTR, kidney transplant recipient; Mean Amp, mean signal amplitude (ion counts); N, number of patients; SD, standard deviation. | | | | | | | | | | | | | | | | | | | | |
| ‡ Peptide identification number. | | | | | | | | | | | | | | | | | | | | |
| † Lower case p indicates hydroxyproline. | | | | | | | | | | | | | | | | | | | | |
| § COL1A1, Collagen α-1(I) chain; COL1A2, Collagen α-2(I) chain; COL2A1, Collagen α-1(II) chain COL3A1, Collagen α-1(III) chain; COL4A2, Collagen α-2(IV) chain; COL4A5, Collagen α-5(IV) chain; EMID1, EMI domain containing 1; GSN, Gelsolin. | | | | | | | | | | | | | | | | | | | | |
| *** AA position according to UniProt Knowledge Base numbering | | | | | | | | | | | | | | | | | | | | |
